# Development and validation of automated methods for COVID-19 PCR MasterMix preparation

**DOI:** 10.1101/2023.10.04.23296537

**Authors:** Giorgio Fedele, Graham Hill, Amelia Sweetford, Suki Lee, Bobby Yau, Domenico R. Caputo, Denise Grovewood, Rowda Dahir, Paula Esquivias Ruiz-Dana, Anika Wisniewska, Anna Di Biase, Miles Gibson, Benita Percival, Stefan Grujic, Donald P. Fraser

## Abstract

Polymerase Chain Reaction (PCR)-based assays were widely deployed during the SARS-CoV-2 pandemic for population-scale testing. High-throughput molecular diagnostic laboratories required a high degree of process automation to cope with huge testing demand, fast turn-around times and quality requirements. However, the critical step of preparing a PCR MasterMix has often been neglected by process developers and optimisers, and is largely dependent upon operator skill for the manual pipetting of reagents to construct the PCR MasterMix. Dependence on manual procedures introduces variation, inconsistency, wastage and potentially risks data integrity. To address this issue, we developed a liquid-handler based solution for automated, traceable and compliant PCR MasterMix preparation. Here, we show that a fully automated PCR MasterMix protocol can substitute manual pipetting, without affecting clinical calling, accuracy or precision. Ultimately, this method reduced cost-per-test at a high-throughput laboratory by eliminating operator-induced wastage while improving the quality of results.

## 1 Introduction

Nucleic Acid Amplification Tests (NAATs) utilise Polymerase Chain Reaction (PCR) (Saiki et al., 1985; Mullis, 1990) as the underlying technology to detect the presence or absence of specific nucleic acid sequences. Such tests were widely employed at large scales across the world during the COVID-19 pandemic to diagnose symptomatic and asymptomatic patients and track the spread of the SARS-CoV-2 virus. PCR-based assays specifically amplify very small quantities of analyte to measurable levels and when compared to other viral detection technologies, such as antigen testing, are recognised as the gold standard in terms of sensitivity and specificity for COVID-19 diagnostics (Leber et al., 2021).

A critical stage in conducting a PCR reaction is the preparation of the PCR MasterMix (MMIX). Typical 1-step MMIXs for the detection of RNA contain Reverse Transcriptase (RT) enzymes, DNA polymerase enzymes, free deoxynucleotide triphosphates (dNTPs), Taqman probes, primers and Mg^2+^ suspended in a buffer in precise concentrations that are optimised for the application. These reagents might be supplied separately, or part combined in pre-prepared concentrations as a kit for ease of use. Laboratory personnel follow protocols detailed in Instructions For Use (IFU) from the Kit Inserts, or Standard Operating Procedures (SOPs) to prepare the MMIX for use in their laboratory’s workflow.

In high throughput laboratories, which during the COVID-19 pandemic processed thousands of NAATs daily, automation of laboratory workflows was vital to achieve targets for turn-around-times, capacity and standardisation. The development of laboratory automation for PCR testing focussed on the sample preparation, purification, and the dispensing protocols for combining purified nucleic acid material and PCR MMIX in microplate formats for PCR reactions (Courtney and Royall, 2021). Even data analysis was largely automated by software solutions (Van Vooren et al., 2022). However, due to the challenges of developing liquid classes for pipetting small volumes of viscous and saponaceous reagents, alongside the need to calculate the correct volumes of reagent to use for different numbers of tests, PCR MMIX preparation was often validated for routine use as a manual only procedure in spite of its extremely high value and quality-critical nature. In regulated environments, it is important that checks are put in place to ensure that data integrity is preserved, but there are challenges with quality control procedures for PCR MMIX preparation. Options include gravimetric checks (which cannot distinguish between two reagents diluted in the same solvent), testing a sample of the batch (costing both time and reagents) or spectroscopic (requiring a spectrophotometer).

An end-point RT-PCR workflow was employed at large scale during the COVID-19 pandemic in the United Kingdom firstly at UK Biocentre, Milton Keynes and then at the Rosalind Franklin Laboratory, Leamington Spa (RFL) (Figure 1). This workflow differed from the more commonly employed RT-quantitative PCR (RT-qPCR) workflows in the analysis stage, where in end-point PCR there is a single fluorescence read at the end of the PCR reaction, as opposed to once every thermal cycle in qPCR (Roix et al., 2021). The process was validated for manual PCR MMIX preparation, with a gravimetric quality control procedure. In operation, meeting performance targets required large batches of PCR MMIX to be created for estimated test numbers, which led to a system that was prone to both wastage and delays through either over-estimation or staff errors. Furthermore, the gravimetric quality control procedures were both slow and insufficient to differentiate between two of the reagents which were both diluted in Tris-HCL buffer. To curtail wastage of PCR reagents at the RFL we developed, validated and implemented an automated, traceable, and compliant process for the preparation of MMIX. Here, we present the results from our development process and validation.

**Figure 1:**
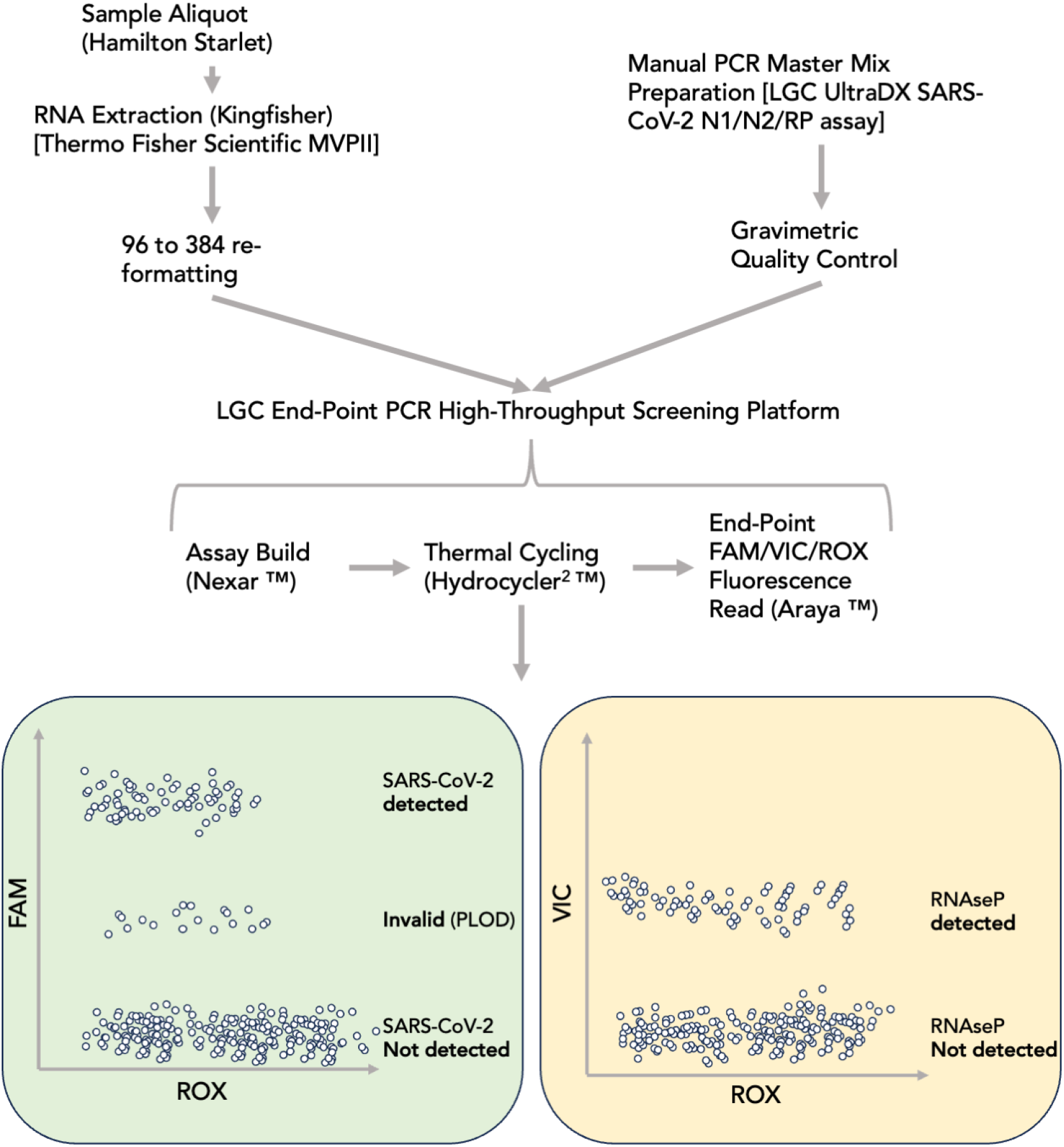
Process flow schematic for SARS-CoV-2 end-point PCR workflow. Samples were purified using the Thermo Fisher Scientific MagMax Viral Pathogen II kit and re-formatted from 96 deep well to 384-well ANSI/SLAS microplates to prepare for assay build in the Nexar Liquid handler 384-well array tape. In parallel, PCR Master Mix was prepared and underwent a Gravimetric Quality Control step before release for a run on the Nexar Liquid Handler. In practice, a run on the LGC End-Point PCR High-Throughput Screening Platform consisted of several (up to 16) 384-well microplates and a batch of PCR Master Mix sufficient for all PCR reactions. The Nexar Liquid Handler first transfers samples with a 384-tip Dispense Pipette from the 384-well microplates to flexible array tape before adding Master Mix to each well of the array tape with a Dispense Jet. The array tape was then sealed and Reverse Transcription and Thermal Cycling were conducted in a Hydrocycler^2^ (HC2) automated water bath. After completion of the PCR reaction, the fluorescence of each well of the array tape was read by the Araya. Collected FAM (SARS-CoV-2 N1 & N2 targets) and VIC (Human RNAseP target) fluorescence data were normalised to ROX loading control and form distinct clusters when plotted.

At the time of writing, COVID-19 has been declared as no longer a Public Health Emergency by the World Health Organisation (WHO, 2023). In the UK, the screening of population samples for SARS-CoV-2 has been largely suspended (Cabinet Office, 2022). Whilst the risk of emergence of a novel immune-escape SARS-CoV-2 variant still exists, the application of high-throughput PCR screening is likely to be limited until the emergence of ’Pathogen X’ (Skyle, 2022). Nevertheless, automated PCR MMIX preparation methods have broad applications across molecular biology and particularly in sequencing pipelines.

## 2 Materials and Methods

### 2.1 Mastermix dispense and PCR conditions

At RFL, SARS-CoV-2 diagnostic NAATs were performed according to the LGC End-Point PCR High-Throughput Screening Platform Instructions-For-Use (IFU) (Figure 1), using manufacturer-supplied equipment (LGC’s Nexar, Hydrocycler^2^ (HC2) & Araya) and LGC’s Biosearch Technologies SARS-CoV-2 ultra-high-throughput End-Point RT-PCR Test reagents (Table 1). Together, this system uses an end-point PCR method where fluorescence values of hydrolysed target and control probes are taken at the end of the PCR reaction. This is unlike quantitative PCR (qPCR) where the fluorescence readings are taken each thermal cycle.

**Table 1:**
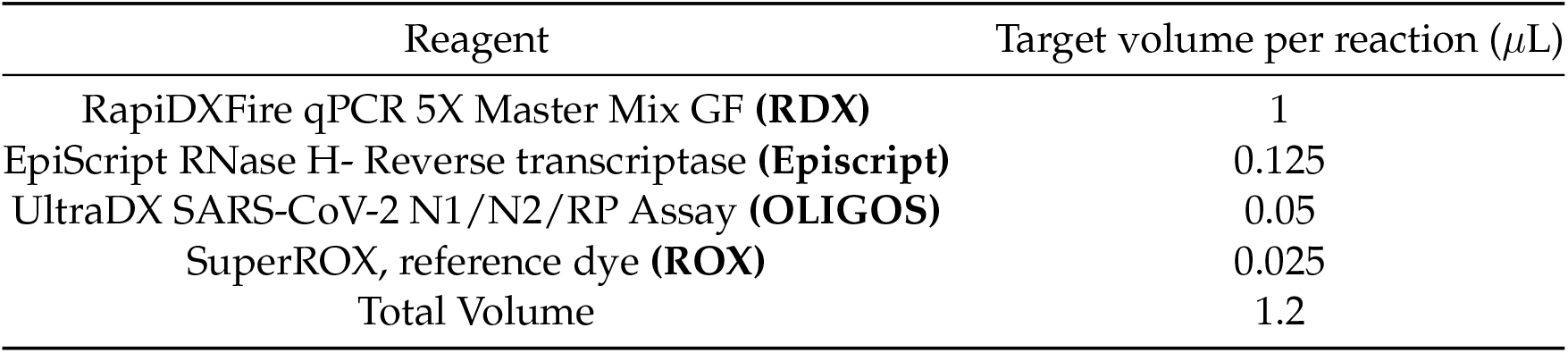
Biosearch Technologies SARS-CoV-2 ultra-high-throughput End-Point RT-PCR Test Reagent List and volumes per reaction.

| Reagent | Target volume per reaction (μL) |
| --- | --- |
| RapiDXFire qPCR 5X Master Mix GF (**RDX**) | 1 |
| EpiScript RNase H- Reverse transcriptase (**Episcript**) | 0.125 |
| UltraDX SARS-CoV-2 N1/N2/RP Assay (**OLIGOS**) | 0.05 |
| SuperROX, reference dye (**ROX**) | 0.025 |
| Total Volume | 1.2 |

Briefly, the Nexar is a modular in-line liquid handling system capable of offering highly scalable throughput owing to its Array Tape (AT) solution, which is a continuous polymer strip that is serially embossed with reaction wells in 384 - well formats.

A CyBio 384 pipette head first transfers the purified RNA (3.8 *µ*l) from a 384-well storage plate into the AT. A Dispense Jet 2.0 (DJ, an 8 tip, single jet solenoid micro-valve system) aspirates *c.* 520 *µ*l of MMIX from a 96 deep-well Nexar Assay Plate (NAP) and then dispenses 1.2 *µ*l of MMIX solution on top of the transferred RNA in the AT. Therefore, one DJ tip is used per array. The AT is sealed with an optically clear seal and transferred to the HC2 for the PCR reaction. The HC2 is an automated water bath system consisting of 3 separate tanks set at different temperatures (50°C, 95°C and 60°C). After the initial Reverse Transcriptase step (50°C for 15 minutes), the AT is cycled 50 times between the 95°C (3 seconds) and 60°C (30 seconds) tanks, giving the denaturation, annealing and extension steps. At the end of the PCR reaction, the AT is manually dried and fed through the Araya, a single optic Photomultiplier Tube (PMT) reader equipped with 3 band pass filters matching the specification of dyes used in the reaction (2.2). A single measure is taken for each well per wavelength and the raw data are corrected using a proprietary deconvolution matrix to correct channel cross-talk. The processed data are exported as a .csv file for analysis.

### 2.2 PCR targets and Result output

LGC’s UltraDX SARS-CoV-2 N1/N2/RP assay targets the SARS-CoV-2 Nucleocapsid (N) gene . The test uses a set of primers designed to anneal to two sections of the N gene (Lu et al., 2020). Fluorescent Taqman probes, consisting of a dye and a quencher, anneal downstream of the primers. Both N gene probes (referred to as N1 and N2) are labelled with a common fluorophore (FAM) and are pooled and detected with the same Araya channel. A third set of primers is designed to anneal to the Human RNase-P (RP) transcript and is used as an internal control. The respective probe is labelled with a VIC fluorophore. ROX dye is also added to reaction both to provide an assay control that can be used to normalise the fluorescence signal and as a quality control step to ensure correct dispensing and addition of the master-mix.

### 2.3 Liquid Handlers

This study compared two liquid handler platforms to manual MMIX preparation, the SPT Labtech Dragonfly Discovery (DF) and the Hamilton StarLet (SL). Methods for creating and dispensing MMIX in NAP were developed for both and are available for download from `https: //github.com/Donald-OMIX/Liquid_Handler_Methods.git`. A Hamilton StarLet fit-ted with a 96-channel pipetting head was used to re-format 96-well plates to 384-format where required.

### 2.4 RNA Purification

Viral RNA was extracted from control materials and patient swabs following the MagMAX Viral/Pathogen II (MVP II) Nucleic Acid Isolation Kit IFU on the Kingfisher Flex platform, supplied by Thermo Fisher Scientific. Note that the extraction protocol theoretically results in a concentrating of the RNA material: Assuming 100% efficiency of extraction, material at 1500 copies mL*^-^*^1^ pre-extraction, would become 5000 copies mL*^-^*^1^ post extraction.

### 2.5 Control Material

For pilot experiments, control materials were supplied by Twist Bioscience. The following synthetic RNA controls were used:

- Control 17, P.1, Gamma Variant (10^9^ copies mL*^-^*^1^)

For validation experiments and in routine use, Randox Laboratories Ltd supplied Qnostics, and Seracare (part of the LGC group) supplied AccuPlex control materials.

- Qnostics SARS-CoV-2 Q Control (SCV2QC) (500 copies mL*^-^*^1^)
- Qnostics SARS-CoV-2 Negative Q Control (TMNQC)
- AccuPlex SARS-CoV-2 Custom Control (1500 copies mL*^-^*^1^)
- AccuPlex SARS-CoV-2 Control (100,000 copies mL*^-^*^1^)

### 2.6 Pilot Experiment Protocol

Twist Gamma variant at 10^9^ copies mL*^-^*^1^ was diluted to 5000, 1500 and 500 copies mL*^-^*^1^ in Elution Buffer from the MVP II kit. Each dilution was then manually dispensed into separate 96 Deep-Well Kingfisher Plates (DWP). The three DWPs were re-formatted alongside an Elution Buffer only DWP to a 384-format plate. It is worth noting that these concentrations are representative of post-extraction concentrations as opposed to the concentrations of crude samples. The 384-format plate was tested twice, firstly with manually-made MMIX, and secondly with the automated MMIX to allow for direct comparison. For each individual experiment, firstly the manual-made MMIX was prepared followed by automated MMIX preparation. As a result of instrument availability while sharing equipment and space with a live operation, within experiments, the Nexars and HC2s were consistent, but between experiments they differed.

Each array was analysed according to standard operational procedures (SOPs) and their standard acceptance criteria. ROX criteria for validity of results is as follows:

- 1600 *<* ROX mean relative fluorescence units (RFU) *<* 4800
- ROX coefficient of variation (CV) % ≤ 20

Dyes were normalised to the ROX RFU, *i.e.* normalised FAM (n FAM) = FAM RFU/ROX RFU, and normalised VIC (nVIC) = VIC RFU/ROX RFU. The resulting normalised values were used for clinical calling whereby, n FAM *≥* 9 is “Detected”, 4 ≤ n FAM *<* 9 is “Inconclusive”, and n FAM *<* 4 is “Not Detected”. Note that as the positive material was a synthetic COVID RNA construct, no RNAse-P signal was detected.

Positive Percent of Agreement (PPA), Negative Percent of Agreement (NPA), Overall Percent of Agreement (OPA) and Cohen’s Kappa were calculated. The 500 copies mL*^-^*^1^ dilution was excluded from these analyses as this concentration falls well below the 95% Lower Limit of Detection (LLoD95), which is the concentration where there is a 95% chance of detection. 500 copies mL*^-^*^1^ post-extraction corresponds to 150 copies mL*^-^*^1^ crude sample, whereas the manufacturer and laboratory-confirmed LLoD95 was *c.* 800 -900 copies mL*^-^*^1^. The 1500 copies mL*^-^*^1^ (450 copies mL*^-^*^1^ crude sample equivalent) was included to stress-test the comparison.

### 2.7 Validation Experiment Protocol

The Validation Protocol was organised around three major experiments:

1. ROX distribution and overall performance of the method, as well as cleanliness of the liquid handler. The objectives of this experiment were to provide confirmation that the equipment:

- Transferred correct volumes of reagent from the source vessels to the destination plate, by comparing ROX distribution from automated MMIX across different arrays.
- Runs without error.
- Was free from contamination (RNA/DNA) that might interfere with the subsequent Validation processes.

No COVID positive material was run, only fresh Elution Buffer, therefore any contamination due to the presence of SARS-CoV-2 RNA/DNA would be revealed after amplification.
2. ePCR Sensitivity. The objective of this experiment was to determine if the automated workflow gave equivalent performance to that of the previously validated Manual process through direct comparison with common MMIX reagents and reference materials. A small study was performed using synthetic SARS-CoV-2 material (Accuplex 100k copies mL*^-^*^1^) at 8 different concentrations (ranging from 2526 to 0 copies mL*^-^*^1^ to determine Analytical Sensitivity. The LLoD95 was calculated using logit analysis. This Analytical Sensitivity represents the smallest amount of substance in a sample that can accurately be measured by an assay and is also referred to as the Lower Limit of Detection (LLoD). It is the lowest concentration of target in a specimen that can be consistently detected at an arbitrarily chosen rate (in this case 95%, LLoD95). Acceptance criteria for LLoD95 was ≤ 1000 copies mL*^-^*^1^, taken from the Target Product Profile for SARS-CoV-2 diagnostics (Cabinet Office, 2020).
3. Concordance. A concordance study between Manual and Automated MMIX preparation was performed using patient samples to determine Diagnostic Sensitivity and Specificity. A Cohen’s Kappa analysis compared the alignment of positive and negative diagnostic calls. Acceptance criteria: essential that the PPA, NPA and OPA are *≥* 95%. Rates *<* 95% would require investigation and root cause analysis. In addition, the Cohen’s Kappa should report a value greater than 0.95.

## 3 Results

### 3.1 Qualitative Evaluation of Liquid Handler Platforms

We began by comparing the liquid handlers that we had in our possession. There were two clear front-runners for this project, the SPT LabTech Dragonfly Discovery (DF); and the Hamilton StarLet (SL) (Table 2). On the basis of its perceived ease-of-use, relatively low complexity, positive displacement pipetting performance, and dispense speed; we opted in the first instance to develop the DF platform for automated MMIX preparation.

**Table 2:**
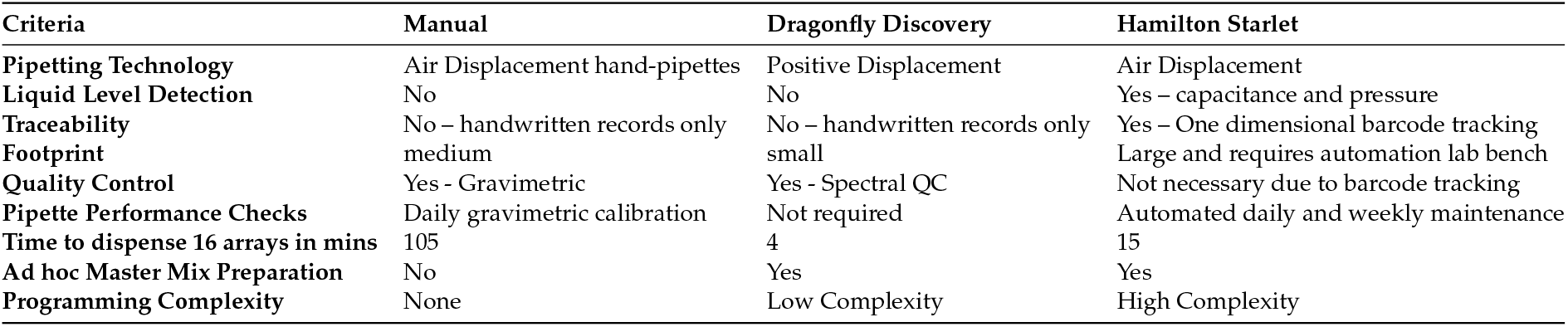
Qualitative comparison of MMIX preparation platforms.

| Criteria | Manual | Dragonfly Discovery | Hamilton Starlet |
| --- | --- | --- | --- |
| Pipetting Technology | Air Displacement hand-pipettes | Positive Displacement | Air Displacement |
| Liquid Level Detection | No | No | Yes – capacitance and pressure |
| Traceability | No – handwritten records only | No – handwritten records only | Yes – One dimensional barcode tracking |
| Footprint | medium | small | Large and requires automation lab bench |
| Quality Control | Yes - Gravimetric | Yes - Spectral QC | Not necessary due to barcode tracking |
| Pipette Performance Checks | Daily gravimetric calibration | Not required | Automated daily and weekly maintenance |
| Time to dispense 16 arrays in mins | 105 | 4 | 15 |
| Ad hoc Master Mix Preparation | No | Yes | Yes |
| Programming Complexity | None | Low Complexity | High Complexity |

### 3.2 Overview of the Dragonfly Discovery method for preparation and dispense of MMIX

The method for the preparation and dispense of the MMIX using the SPT Dragonfly Discovery instrument (DF) uses 5 reservoirs and syringes and a 96-Deep Well Nexar assay plate (NAP). The system aspirates all the required reagents at once, but dispenses them in individual wells, thus effectively creating different MMIX in each well. The reagents are dispensed in order according to the SOP, that is RDX, ROX, OLIGO and Episcript (refer to Table 1 for reagent list). This specific order was enforced by modification of the dispense pattern to typewriter, as opposed to serpentine, and the aspiration location of the reagents. Two separate dispense layers were created: The first dispenses RDX, ROX and OLIGOS, while the second dispenses only Episcript. The rationale for dividing the dispense into two stages is that Episcript needs to be kept cold until its addition to the mix, thus instead of aliquoting all the reagents into the reservoir at the beginning of the run, the operator dispenses Episcript only after the first layer has been completed. After the first layer has run, the machine stops and asks for manual intervention. At that point the operator adds Episcript in the dedicated reservoir and resumes the run. The two layers can be run using the “Sequence Launcher” function. For a 16 array run, the DF will take *c.* 4 minutes from start to finish.

- Positions 7 and 8 of the reservoir tray were reserved for two standard reservoirs filled with up to 4.2 mL of RDX each to account for dead volume (syringe maximum capacity is 4 mL).
- Position 6 was dedicated to ROX, with 230 *µ*L dispensed into a low-dead-volume (LDV) reservoir.
- Position 5 held OLIGO, with up to 440 *µ*L in a LDV reservoir.
- Position 4 was reserved for Episcript, up to 1.1 mL in a LDV reservoir.

Channel 4 was configured for minimum speed of aspiration and dispense of Episcript, which is highly viscous. After dispense, the MMIX plate was sealed and shaken for 30 seconds at 900 rpm on an Eppendorf Thermomixer to ensure resuspension and mixing of reagents.

### 3.3 ROX RFU is shifted in Dragonfly-prepared MMIX

Using the above DF configuration (3.2), automated MMIX ROX statistics were within acceptance criteria (Table 3). However, the distributions of ROX RFU and FAM RFU were both left-shifted in the DF-prepared MMIX compared to Manual (Figure 2A & B). Lower ROX values could lead to a discrepancy in diagnostic calling as low ROX RFU will push n FAM values higher (Figure 2C & D).

**Figure 2:**
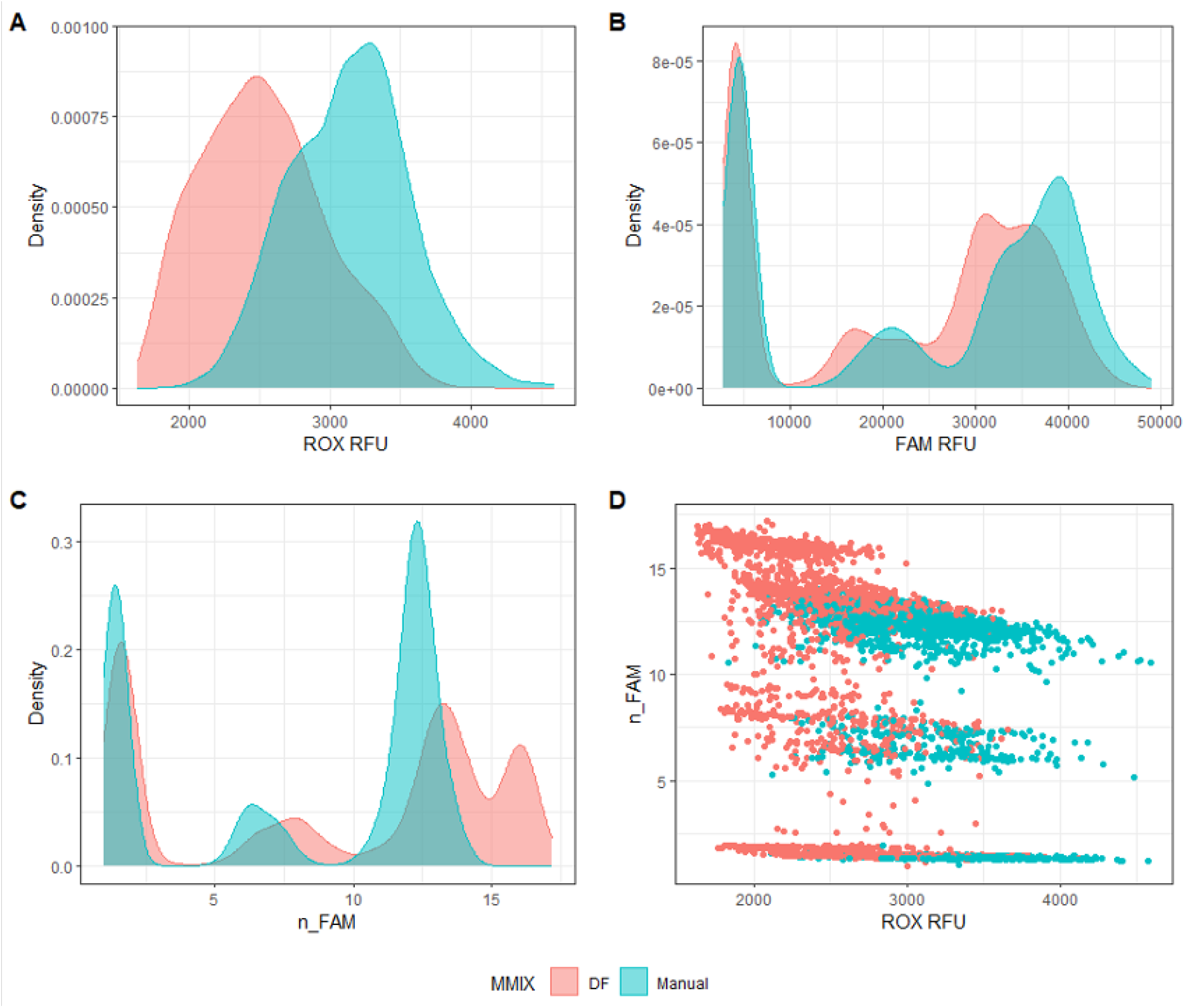
Dragonfly Discovery-Prepared MMIX compared with Manually-Prepared MMIX. End-point RFU data from PCR reactions using dilutions of synthetic RNA positive control material (Twist) at and around the limit of detection. A) ROX RFU kernel distribution. B) Distribution of FAM RFU values. C) Distribution of n FAM values. D) Scatterplot n FAM vs ROX RFU. Data are pooled from six 384-well arrays for both conditions.

**Table 3:** ROX Statistics of Dragonfly and Manual-prepared MMIX. N = 384.

| MMIX Preparation Method | Tape Id | Mean | SD | CV (%) |
| --- | --- | --- | --- | --- |
| Dragonfly | 738732 | 2359.7 | 276.8 | 11.7 |
|  | 738733 | 2060.1 | 282.9 | 13.7 |
|  | 852899 | 3024.8 | 349.4 | 11.6 |
|  | 852901 | 2532.3 | 362.6 | 14.3 |
|  | 852903 | 2623.7 | 369 | 14.1 |
|  | 852905 | 2540.6 | 362.4 | 14.3 |
| Manual | 738734 | 3252.7 | 325 | 10 |
|  | 738735 | 2962.5 | 313.7 | 10.6 |
|  | 852900 | 3057.1 | 423 | 13.8 |
|  | 852902 | 2951.4 | 417.8 | 14.2 |
|  | 852904 | 3200.1 | 394.4 | 12.3 |
|  | 852906 | 3398.7 | 434.5 | 12.8 |

### 3.4 Addition of emulgent (Tween-20) ameliorates ROX distribution

A root-cause-analysis was performed to understand the reasons for the low ROX RFU values observed (3.3). Under dispensing was ruled out as a volumetric check revealed dispense accuracy and precision within our acceptance criteria (data not shown). We postulated that plastic binding might be responsible for the drop in RFU as rhodamine derivatives (such as the ROX fluorophore) adsorb to plastics (Du et al., 2022). We decided to overcome this potential issue by adding an emulgent to ROX, Tween-20. This non-ionic surfactant was normally added to keep the ROX fluorophore in solution, however, for regulatory compliance reasons, it was later removed from the final product by the manufacturer. A preliminary experiment was performed where 0.05% Tween-20 (provided by LGC) was added to a subaliquot of ROX, and the MMIX was then prepared both manually and with the DF, and the experiment described above (3.3) was repeated.

The addition of Tween brought ROX statistics and distribution for both the Manual and DF-prepared MMIX into alignment (Table 4 & Figure 3A). In addition, the FAM distributions between of the Manual and DF-prepared MMIX were also closely aligned (Figure 3B). Correspondingly, they showed overlapping n FAM distributions (3C & D). Tape Id 740959 resulted in a ROX CV % *>* 20. While this figure exceeds the acceptance criteria, the most likely cause of this issue is attributable to a Nexar mis-dispense rather than a DF mis-dispense. This is because the DF aspirates all the required volume for ROX at once. If not enough liquid is present in the reservoir, air will be aspirated instead creating a gap between the plunger and the meniscus, which would only impact the final dispense of a run (Tape Id 740960), rather than the middle dispense.

**Figure 3:**
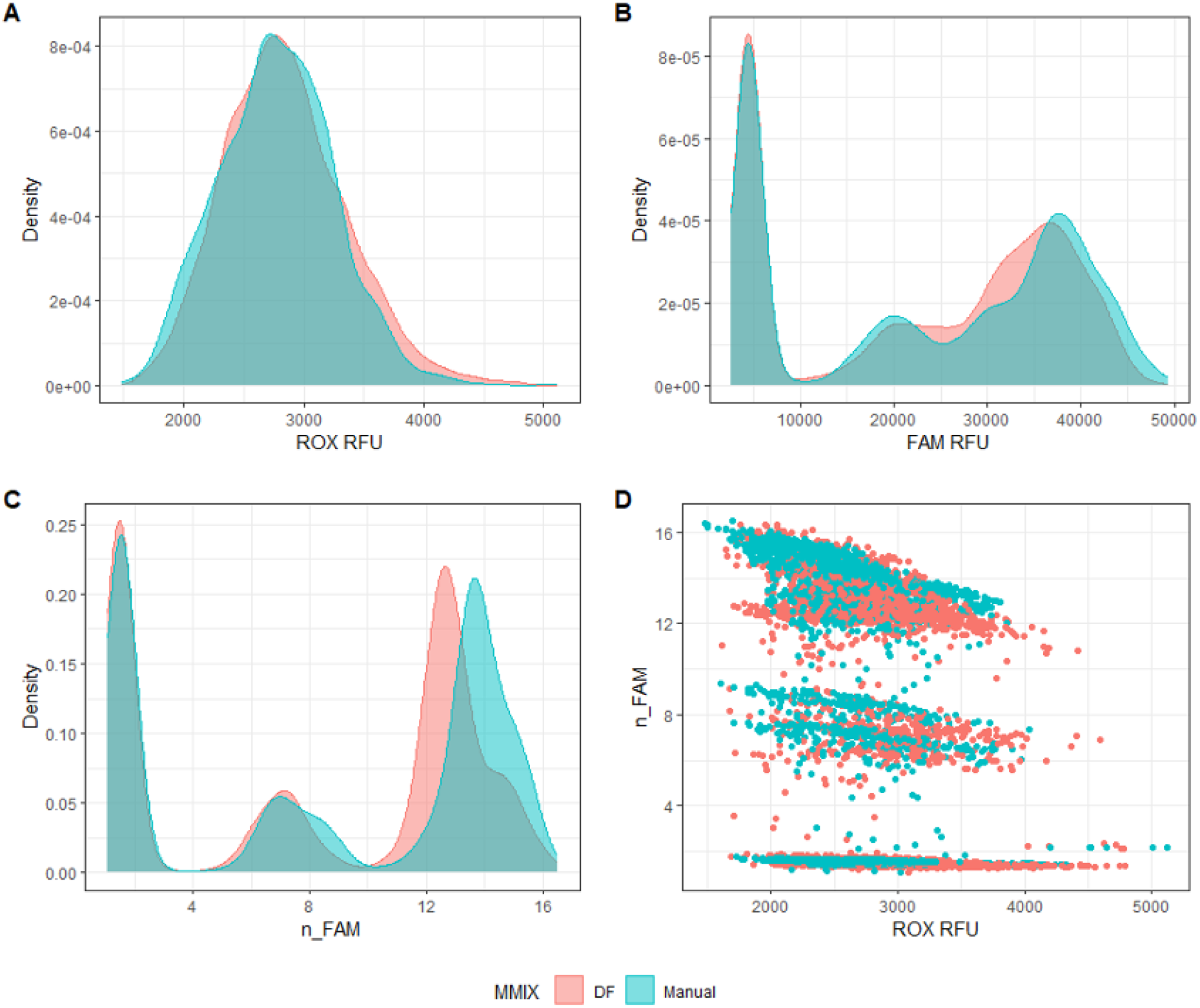
Dragonfly Discovery-Prepared MMIX compared with Manually-Prepared MMIX after addition of 0.05% Tween-20. End-point RFU data from PCR reactions using dilutions of synthetic RNA positive control material (Twist) at and around the limit of detection. A) ROX RFU kernel distribution. B) Distribution of FAM RFU values. C) Distribution of n FAM values.

**Table 4:**
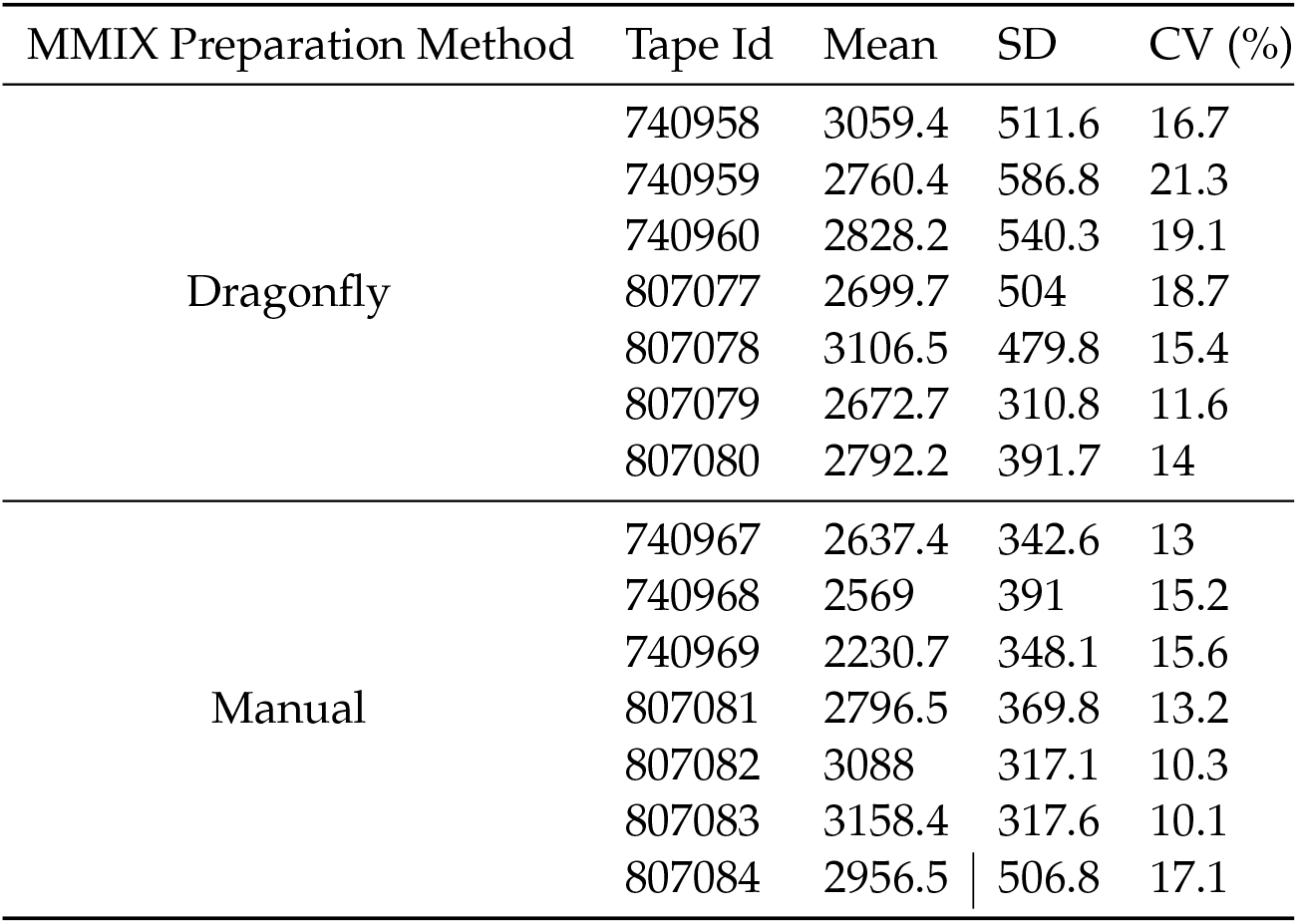
ROX Statistics of Dragonfly and Manual-prepared MMIX with Tween-20 added. N = 384.

| MMIX Preparation Method | Tape Id | Mean | SD | CV (%) |
| --- | --- | --- | --- | --- |
| Dragonfly | 740958 | 3059.4 | 511.6 | 16.7 |
|  | 740959 | 2760.4 | 586.8 | 21.3 |
|  | 740960 | 2828.2 | 540.3 | 19.1 |
|  | 807077 | 2699.7 | 504 | 18.7 |
|  | 807078 | 3106.5 | 479.8 | 15.4 |
|  | 807079 | 2672.7 | 310.8 | 11.6 |
|  | 807080 | 2792.2 | 391.7 | 14 |
| Manual | 740967 | 2637.4 | 342.6 | 13 |
|  | 740968 | 2569 | 391 | 15.2 |
|  | 740969 | 2230.7 | 348.1 | 15.6 |
|  | 807081 | 2796.5 | 369.8 | 13.2 |
|  | 807082 | 3088 | 317.1 | 10.3 |
|  | 807083 | 3158.4 | 317.6 | 10.1 |
|  | 807084 | 2956.5 | 506.8 | 17.1 |

### 3.5 UV-VIS spectroscopy can be used for quality control of prepared MMIX

At RFL, before usage in routine clinical analysis, MMIX was subjected to a gravimetric quality control check (QC). While this method is simple and does not required specialised equipment (besides a balance), it is not suitable for detecting a common operator mistake: the swapping of ROX and OLIGO blend volumes. The reason being that both reagents are diluted in the same buffer (Tris-HCl based buffer). Hence, their mass:volume is near identical. Such an error would lead to substantial differences in the clinical calling, as the ratio between the normaliser (ROX) and the real signal (FAM/VIC probe blend) is incorrect. To overcome this issue, we developed a D) Scatterplot n FAM vs ROX RFU. Data are pooled from seven 384-well arrays for both conditions. new method whereby the absorbance of the MMIX is measured at defined wavelengths, namely 502, 550 and 585 nm, which are the peak absorbtion wavelengths of FAM, VIC and ROX. By calculating a ratio between the different peaks it was therefore possible to define a range within which a MMIX is deemed correct (3 SD from the mean). MMIX created either manually or with the DF were compared and common errors in MMIX preparation were simulated (*e.g.* swapping the volumes of ROX and OLIGOs).

For this experiment, we used a PheraSTAR plate reader, on loan from BMG LABTECH. 10 *µ*L of MMIX (both correct and swapped) were dispensed into 4 wells of a flat bottom plate (Corning UV micro-star LV) and a spectral scan was performed. As depicted in figure 4, the absorption spectral scan (ABS) revealed clear defined peaks at the predicted wavelengths for the normal and swapped MMIX. By calculating the ratio between FAM/ROX and VIC/ROX it was possible to observe a clear cluster separation between the two MMIXs (Figure 5).

**Figure 4:**
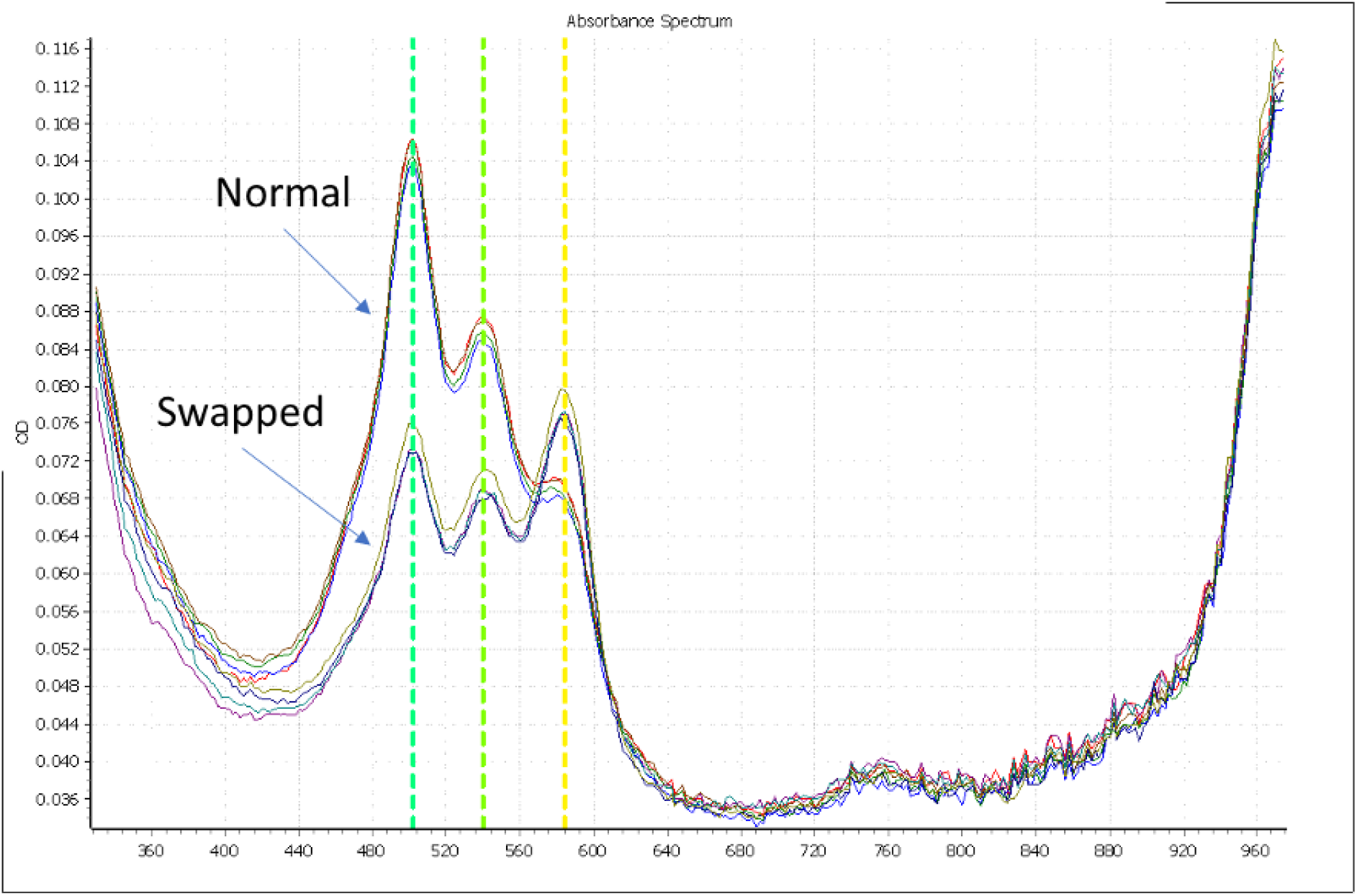
ABS scan, comparing the absorption spectra of “Normal” MMIX, and a “Swapped” MMIX, where the volumes of the ROX and OLIGO are exchanged. N = 4.

**Figure 5:**
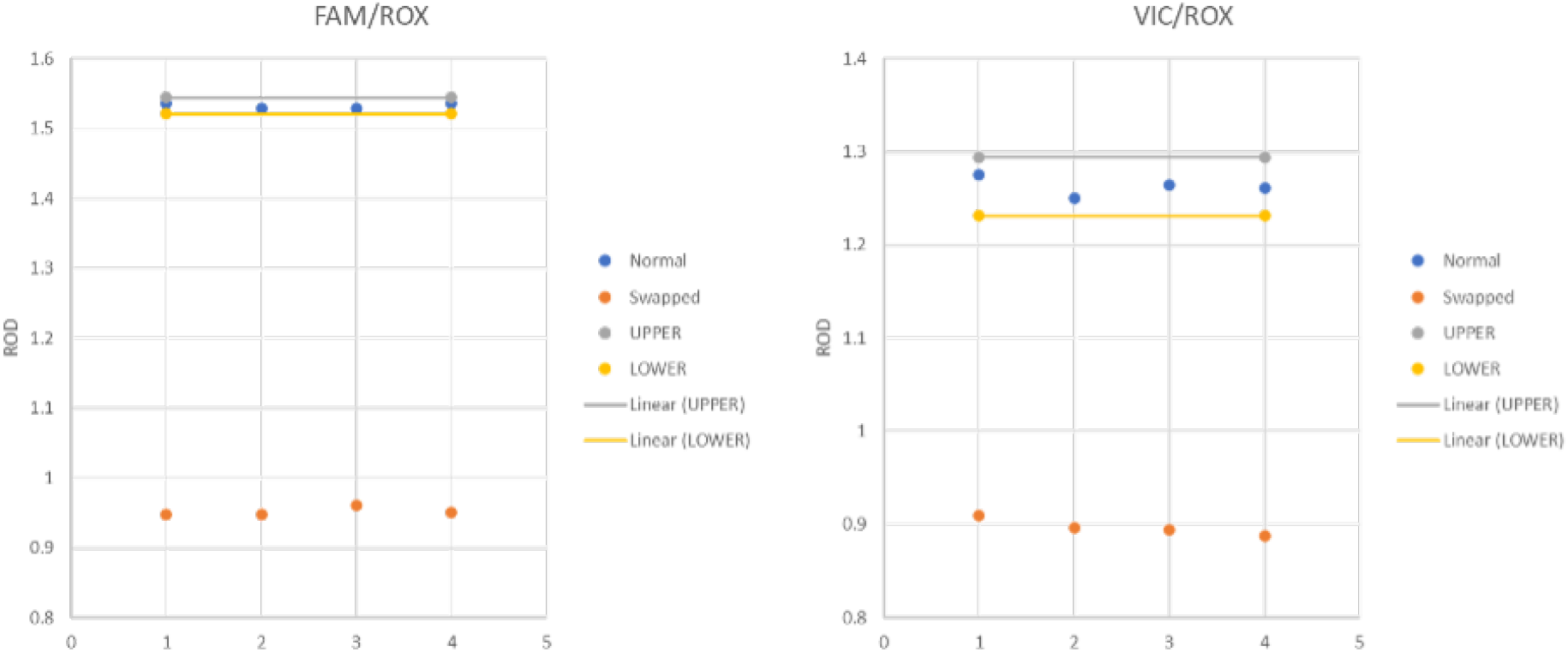
Ratio of FAM/ROX and VIC/ROX, with upper and lower limits defined as *±* 3 SD from the mean. N = 4.

To investigate further the sensitivity of the spectrometric QC step, we tested several other MMIX errors, which could occur (Figure 6A). A comparison between DF and Manual-prepared MMIX was also performed. Manually-prepared and DF-prepared MMIX clusters overlapped (Figure 6B), again confirming the ability of the DF to produce MMIX of a quality comparable to an experienced operator.

**Figure 6:**
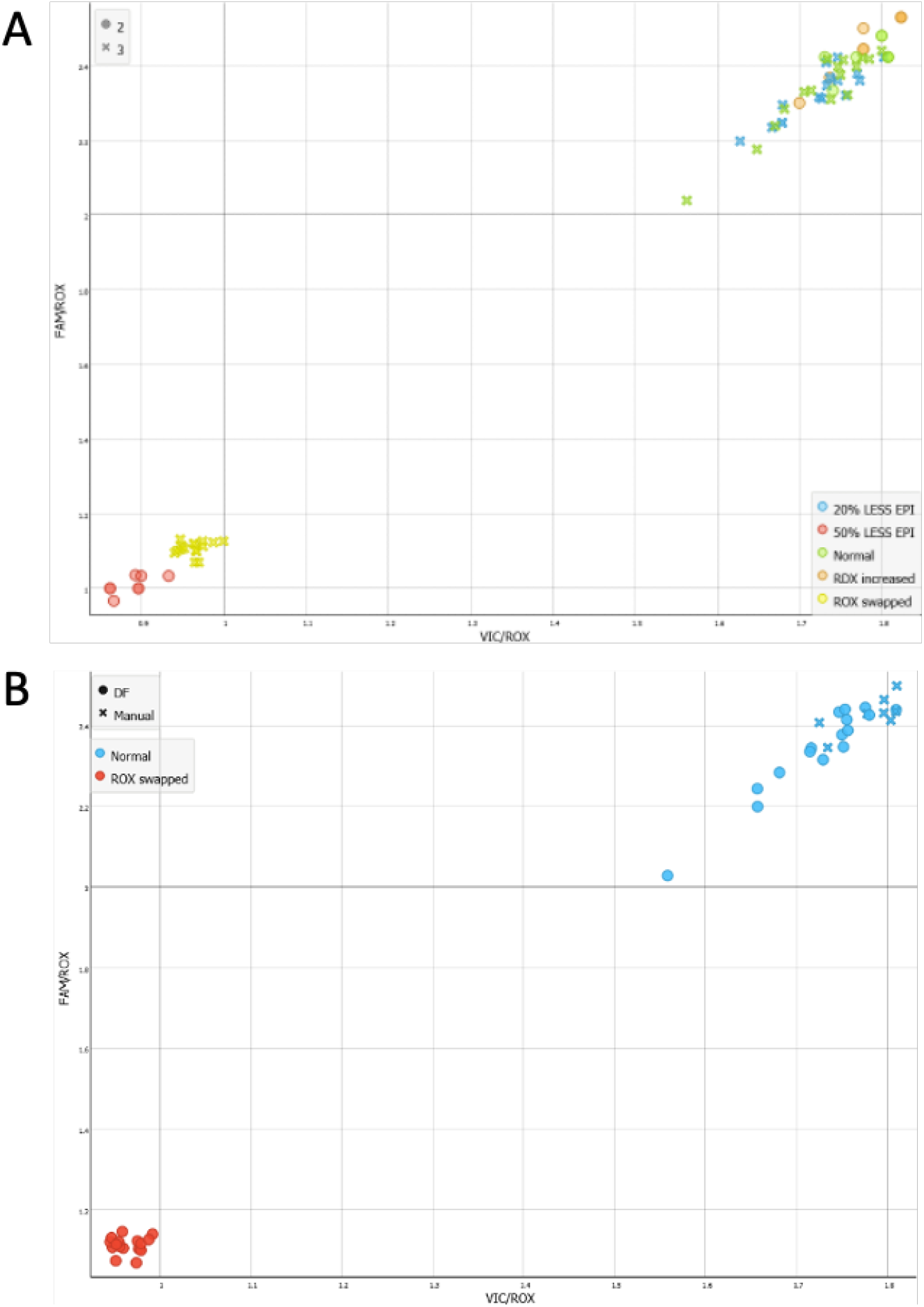
Clustering of MMIX Absorption spectra. A) Simulated Common Errors. B) Comparison of DF-prepared MMIX, Manually-prepared MMIX, and with ROX and OLIGO swapped. Note EPI = Episcript.

These data together suggest that the spectrometric QC assay could be a highly sensitive method for detecting incorrectly prepared MMIX, Particularly for cases that could not be otherwise detected gravimetrically. Similarly, other “errors” like a 50% reduction in volume of Episcript can also be detected. However, despite the sensitivity of the instrument, small variations in the Episcript volume (20% reduction) could not be detected spectro-metrically using our method. There is the potential for use of gravimetric detection, as a variation of 20% will shift the weight of MMIX by 2.1%, raising the possibility of using both QC methods in concert as a more robust detection system, should the application require it.

In spite of its equivalent performance to a skilled manual operator, we were unable to take the DF to a routine operational environment as to do so would require the spectroscopic QC, and unfortunately the PheraSTAR plate reader loan unit had to be returned. We therefore proceeded to develop our other liquid handler platform, the Hamilton StarLet. Pivoting to an alternate platform presented an opportunity to utilise the pipette log, liquid level detection and tracking features of the StarLet platform, which together could bypass the requirement for the QC step entirely.

### 3.6 Overview of the StarLet method for preparation and dispense of MMIX

The StarLet method was designed and programmed to maximise reagent recovery in the event of the method aborting due to (*e.g.*) an aliquot running out, or the operator loading labware or reagents incorrectly. To achieve this, liquid level detection and barcode tracking of all reagents containers was implemented. The logic of the method was developed to minimise the risk of wastage of expensive reagents while respecting the requirements for keeping the Episcript cold (Algorithm 1). Briefly, RDX, ROX and OLIGO are not mixed until the method confirms that all three reagents are available in sufficient quantity. RDX is dispensed first as it is added in bulk, and if left in the tip while the other reagents are aspirated would begin to drip. The method will then request Episcript, and if sufficient quantity confirmed, the method will proceed with to combine the Episcript with the mixed reagents.

### 3.7 The Hamilton StarLet provides equivalent pipetting performance to a manual operator

Comparison of manually-prepared MMIX with Hamilton StarLet (SL)-prepared MMIX showed a high degree of overlap for both ROX, and FAM RFU distributions (Figure 7). Furthermore,

**Figure 7:**
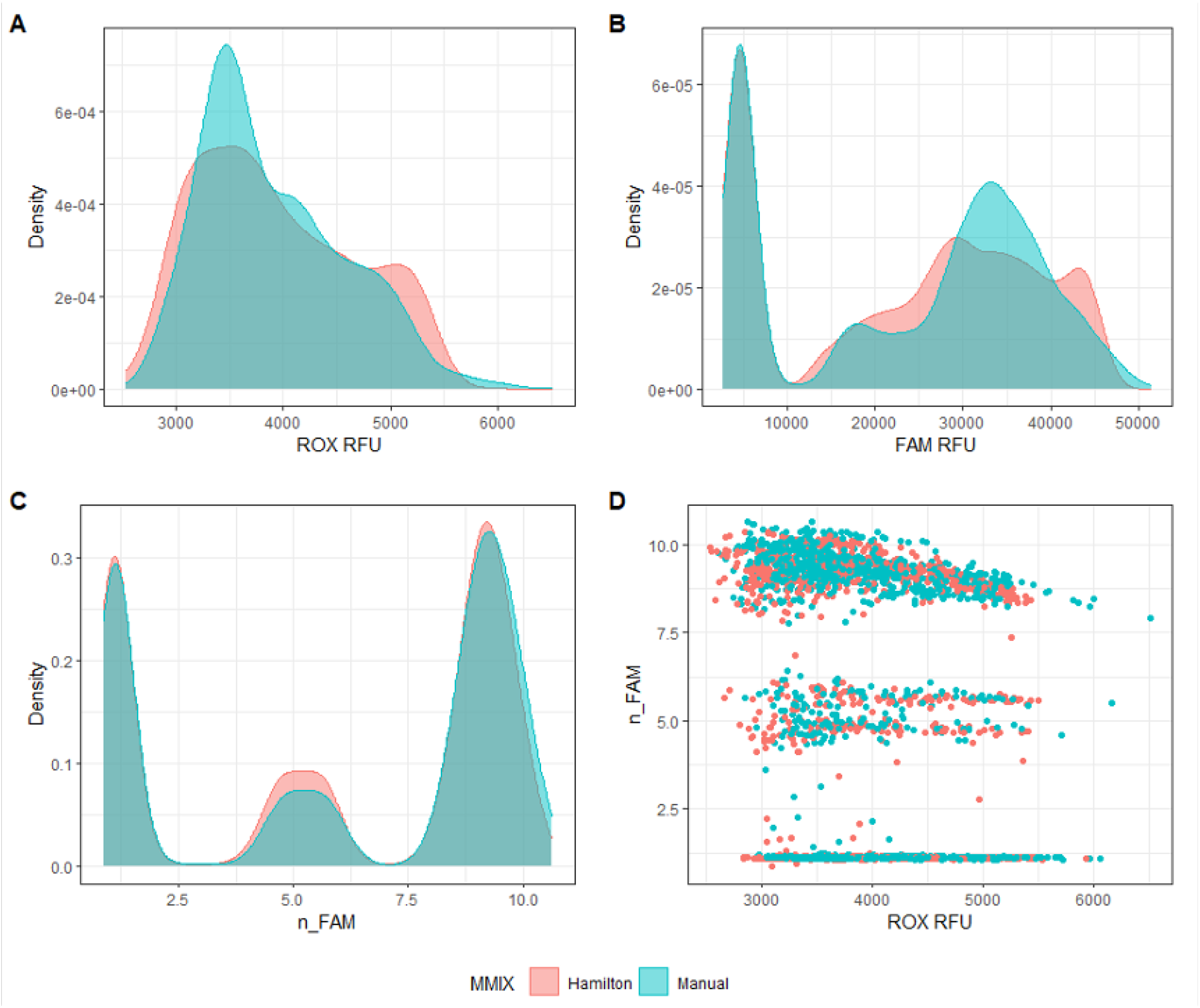
StarLet-Prepared MMIX compared with Manually-Prepared MMIX. End-point RFU data from PCR reactions using dilutions of synthetic RNA positive control material (Twist) at and around the limit of detection. A) ROX RFU kernel distribution B) FAM RFU kernel distribution. C) n FAM kernel distribution. D) Scatterplot n FAM vs ROX RFU. Data are pooled from three 384-well arrays for both conditions.

**Algorithm 1.**
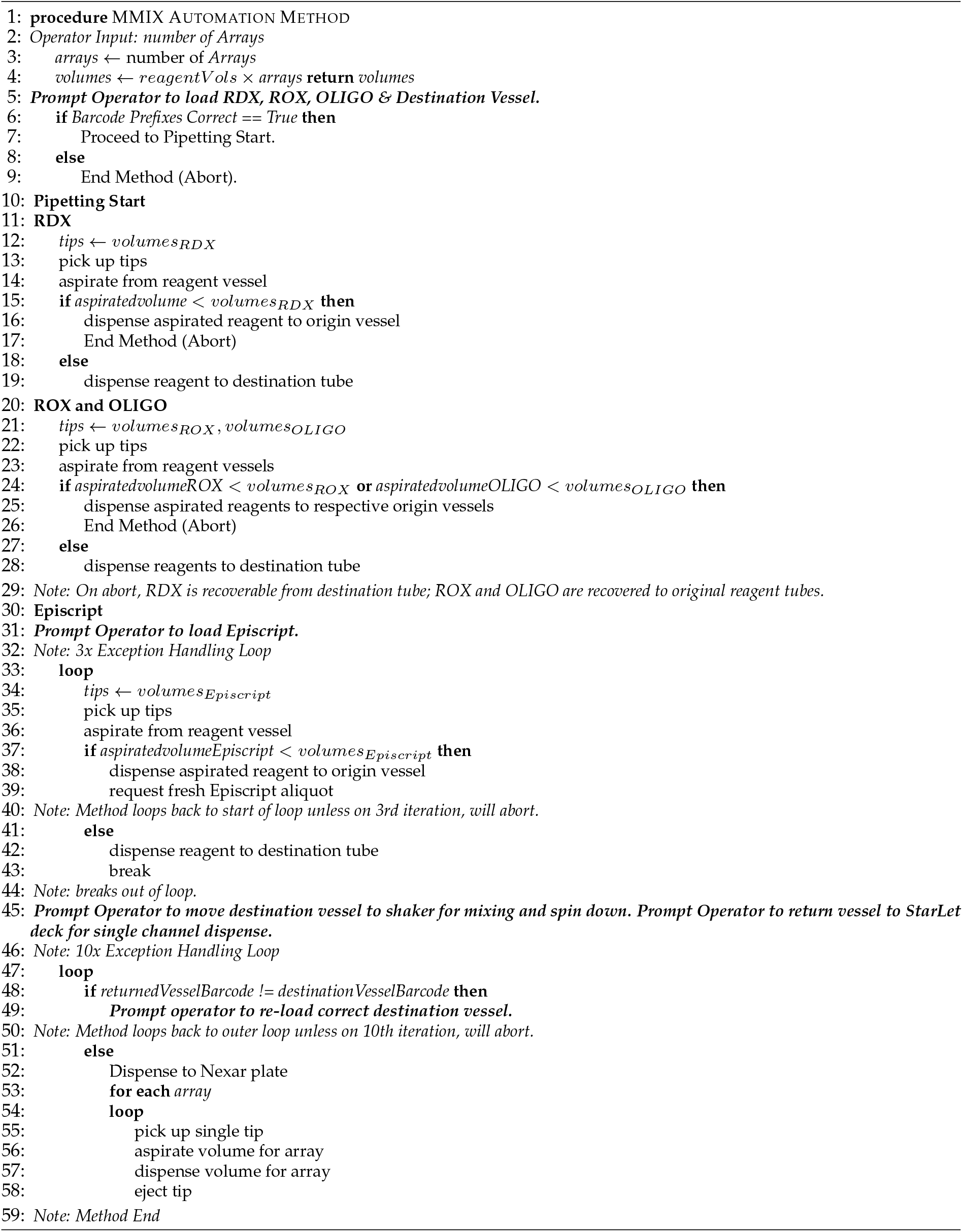
Hamilton StarLet MMIX Automation Method Pseudocode.

ROX statistics were within acceptance criteria (Table 5). Based on the promise showed by these data, the method was validated for routine processing of clinical samples.

**Table 5:** ROX Statistics of Hamilton StarLet and Manual-prepared MMIX. N = 384.

| MMIX Preparation Method | Tape Id | Mean | SD | CV (%) |
| --- | --- | --- | --- | --- |
| Hamilton | 596217 | 4623.4 | 587.8 | 12.7 |
|  | 596219 | 3913.9 | 510 | 13 |
|  | 596221 | 3284 | 361.2 | 11 |
| Manual | 596218 | 4440.7 | 638.6 | 14.4 |
|  | 596220 | 3787.1 | 597.1 | 15.8 |
|  | 596222 | 3502.9 | 356.7 | 10.2 |

### 3.8 Automated preparation of MMIX by the Hamilton StarLet was validated for routine diagnostic use

The first validation experiment sought to assess the overall performance of the method through a formal assessment of ROX statistics (Table 6). Note that the first dispensed array (567099) showed higher ROX CV % (19%) compared to the other dispensed arrays. Given that the MMIX is the same for the first 4 arrays (567099 - 567102), it can be excluded that the issue is related to the MMIX preparation, but rather a lower dispensing performance of the DJ tip on the Nexar, which was a known and recurring issue.

**Table 6:** ROX Statistics for formal validation experiment of StarLet method. N = 384.

| MMIX Preparation Method | Tape Id | Mean | SD | CV (%) |
| --- | --- | --- | --- | --- |
| Hamilton | 567099 | 3095.2 | 586.8 | 19 |
|  | 567100 | 3008.2 | 438.3 | 14.6 |
|  | 567101 | 3346.2 | 421.7 | 12.6 |
|  | 567102 | 3673.8 | 517 | 14.1 |
| Manual | 567103 | 3106.6 | 396.8 | 12.8 |
|  | 567104 | 3308.4 | 354.6 | 10.7 |
|  | 567105 | 3656.5 | 301.9 | 8.3 |
|  | 567106 | 2861.5 | 324.8 | 11.4 |

The distribution of ROX and FAM between the two MMIX was highly comparable (Figure 8A & B). The observation that there were no positive wells and a total of 3 inconclusive wells (equal to 0.1% of total wells) confirmed that the instruments and methods were contaminant free over the 8 examined arrays (Figure 8C & D).

**Figure 8:**
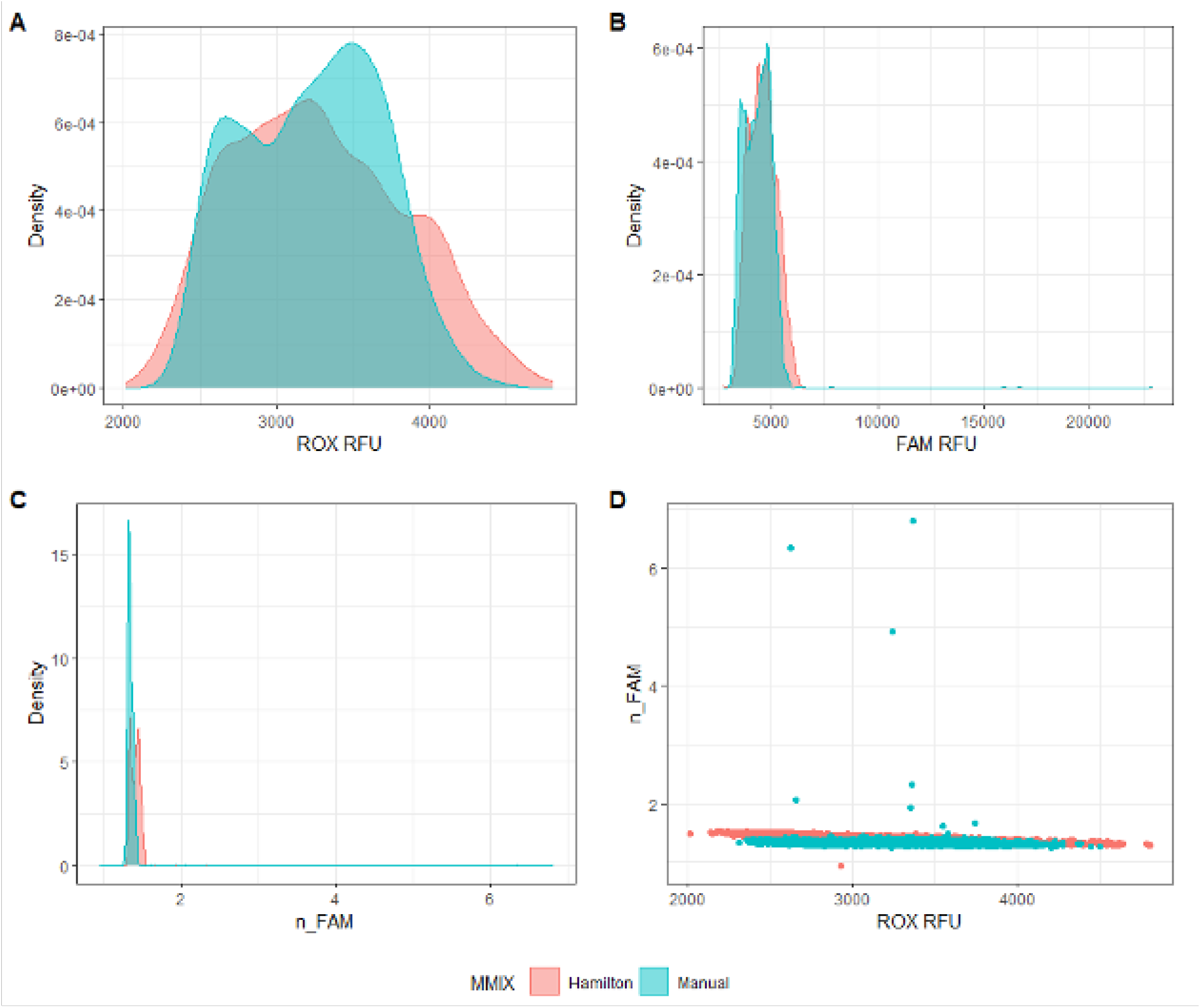
StarLet-Prepared MMIX compared with Manually-Prepared MMIX. End-point RFU data from PCR without inclusion of positive control material to check for contamination. A) ROX RFU kernel distribution B) FAM RFU kernel distribution. C) n FAM kernel distribution. D) Scatterplot n FAM vs ROX RFU. Data are pooled from four 384-well arrays for both conditions.

The second validation experiment assessed the analytical sensitivity of the ePCR assay comparing MMIX prepared with the automated StarLet method and manual workflows. A dilution series of synthetic control material (AccuPlex SARS-CoV-2 100,000 copies mL*^-^*^1^) was prepared (n = 96 for each concentration). The dilution series was tested against both automated and manually-prepared MMIX. The experiment was repeated three times and a logit analysis (Figure 9) was performed to determine the analytical sensitivity (LLoD95) (Table 7). The analytical sensitivity for both MMIX preparation methods met the acceptance criteria of ≤ 1000 copies mL*^-^*^1^, with 0 false positives. In addition, all 12 arrays passed the ROX volumetric dye acceptance criteria (CV% ≤ 20%) (Table 8) and the overall distributions of ROX between automated and manually-prepared MMIX were highly comparable (Figure 10A). Despite a slight right-ward shift in FAM distribution (Figure 10B), n FAM was also closely aligned (Figure 10C & D).

**Figure 9:**
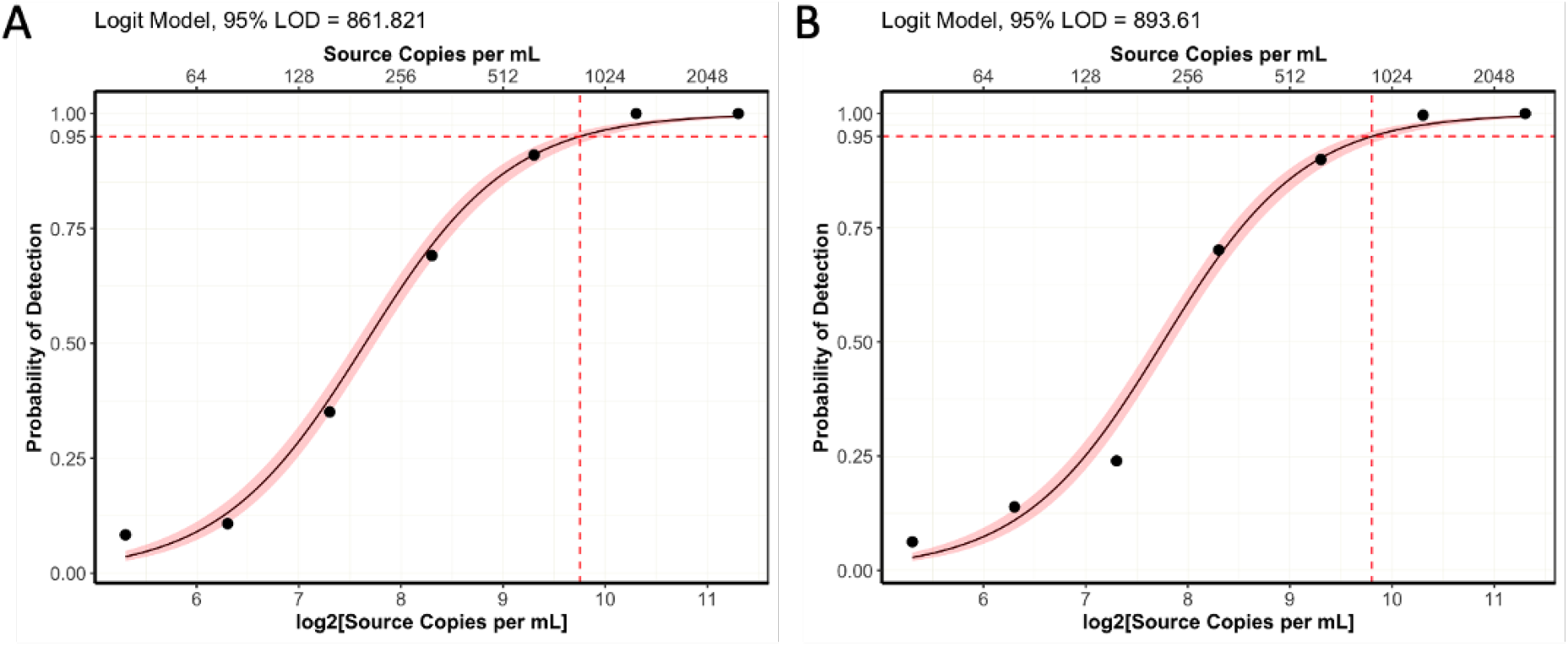
Logit models for analytical sensitivity of A) Manual process B) Automated process. N = 96 per concentration for both conditions. Pink shaded region represents 95% confidence interval.

**Figure 10:**
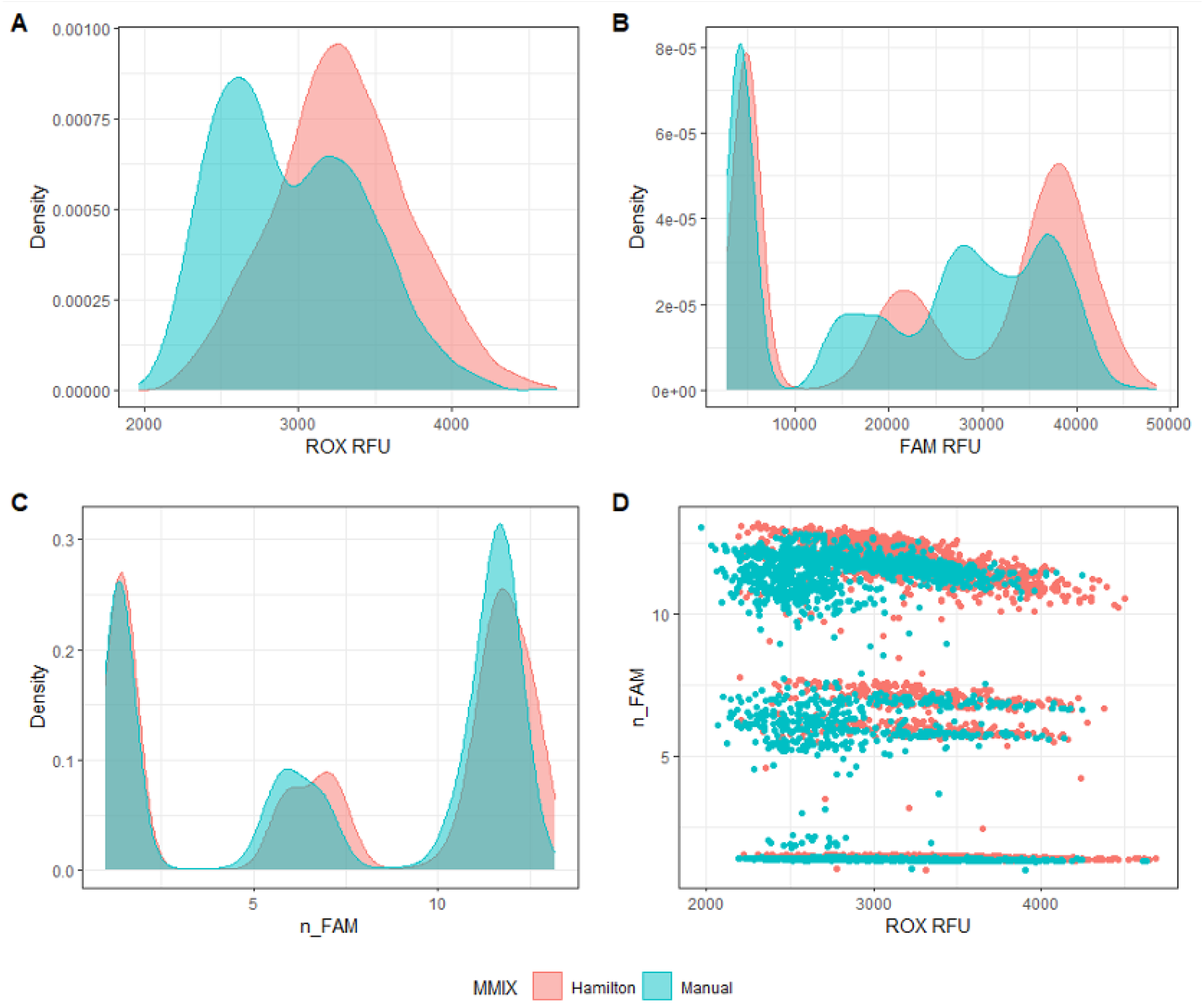
StarLet-Prepared MMIX compared with Manually-Prepared MMIX. End-point RFU data from PCR reactions using dilutions of synthetic RNA positive control material (Seracare AccuPlex) at and around the limit of detection. A) ROX RFU kernel distribution B) FAM RFU kernel distribution. C) n FAM kernel distribution. D) Scatterplot n FAM vs ROX RFU. Data are pooled from six 384-well arrays for both conditions.

**Table 7:** Summary of analytical sensitivity of automated and manually-prepared MMIX.

| Method of MMIX Preparation | Replicate ID | LLoD95 (copies mL<sup>-1</sup>) |
| --- | --- | --- |
| Hamilton | Titre 1 | 931 |
|  | Titre 2 | 808 |
|  | Titre 3 | 935 |
| Manual | Titre 1 | 831 |
|  | Titre 2 | 837 |
|  | Titre 3 | 918 |

**Table 8:** ROX Statistics for ePCR sensitivity experiments. N = 384.

| MMIX Preparation Method | Tape Id | Mean | SD | CV (%) |
| --- | --- | --- | --- | --- |
| Hamilton | 567110 | 3038 | 395.9 | 13 |
|  | 567111 | 3247.4 | 377.1 | 11.6 |
|  | 567112 | 3572.9 | 487.7 | 13.6 |
|  | 567113 | 3342.5 | 419.6 | 12.6 |
|  | 567114 | 3340.9 | 345.2 | 10.3 |
|  | 567115 | 3195.9 | 381.3 | 11.9 |
| Manual | 567116 | 3274.5 | 449.3 | 13.7 |
|  | 567117 | 3122.1 | 575.2 | 18.4 |
|  | 567118 | 3009.5 | 344.4 | 11.4 |
|  | 567119 | 2992.4 | 420.3 | 14 |
|  | 567120 | 2743.9 | 221.7 | 8.1 |
|  | 567121 | 2561.2 | 263.5 | 10.3 |

The final validation experiment sought to determine diagnostic sensitivity through comparison of the automated MMIX procedure with the validated manual procedure using true patient samples. A set of 12 384-well storage plates containing purified patient samples were chosen at random to be first subjected to the validated in-use manual MMIX process, and then by the automated MMIX process.

The performance of the ROX volumetric dye passed acceptance criteria for 23 out of 24 arrays. Tape ID 567232 had a cluster of 6 wells with ROX RFU *>* 5000 RFU. Root-cause-analysis revealed that this array was dispensed by Nexar DJ tip 5, which had previously recorded performance issues. All other wells had ROX RFU within the expected range, therefore excluding these 6 wells from the analysis allowed for the confirmation that the MMIX preparation by the StarLet and the MMIX dispense by the Nexar platform met requirements for diagnostic validity (Table 9).

**Table 9:** ROX statistics of all arrays from the concordance study. N = 384.

| MMIX Preparation Method | Tape Id | Mean | SD | CV (%) |
| --- | --- | --- | --- | --- |
| Hamilton | 567228 | 2683.8 | 405.9 | 15.1 |
|  | 567229 | 3412.3 | 406.5 | 11.9 |
|  | 567230 | 3203.6 | 452.4 | 14.1 |
|  | 567231 | 3110.4 | 329.2 | 10.6 |
|  | 567232 | 3372.4 | 1150.2 | 34.1 |
|  | 567233 | 3159.5 | 328.3 | 10.4 |
|  | 567234 | 3206.8 | 487.6 | 15.2 |
|  | 567235 | 2652.5 | 307.1 | 11.6 |
|  | 567265 | 3193.4 | 419.8 | 13.1 |
|  | 567266 | 3608.3 | 456.6 | 12.7 |
|  | 567267 | 3564.2 | 508 | 14.3 |
|  | 567268 | 3090.8 | 591.9 | 19.2 |
| Manual | 567173 | 2949 | 385.6 | 13.1 |
|  | 567174 | 2585.8 | 334.2 | 12.9 |
|  | 567175 | 2625.3 | 329.8 | 12.6 |
|  | 567176 | 3019.2 | 276.3 | 9.2 |
|  | 567177 | 3003.5 | 487.6 | 16.2 |
|  | 567178 | 3318.9 | 371.6 | 11.2 |
|  | 567179 | 3375.1 | 410.2 | 12.2 |
|  | 567180 | 3000.4 | 493.4 | 16.4 |
|  | 819028 | 3277.2 | 392.3 | 12 |
|  | 819059 | 2710.6 | 297.1 | 11 |
|  | 819061 | 2930.1 | 225.5 | 7.7 |
|  | 819078 | 2654.5 | 348.5 | 13.1 |

ROX and n FAM distribution were again very similar between automated and manually prepared MMIX methods (Figure 11A & B). Correlation between n FAM vs ROX and n VIC vs ROX also showed overlapping distributions between the two MMIXs (Figure 11C & D). A Cohen’s Kappa analysis of the diagnostic results showed close correlation and agreement on results (*≥* 95%) between the two MMIX preparation methods when testing patient samples (Table 10 & Table 11).

**Figure 11:**
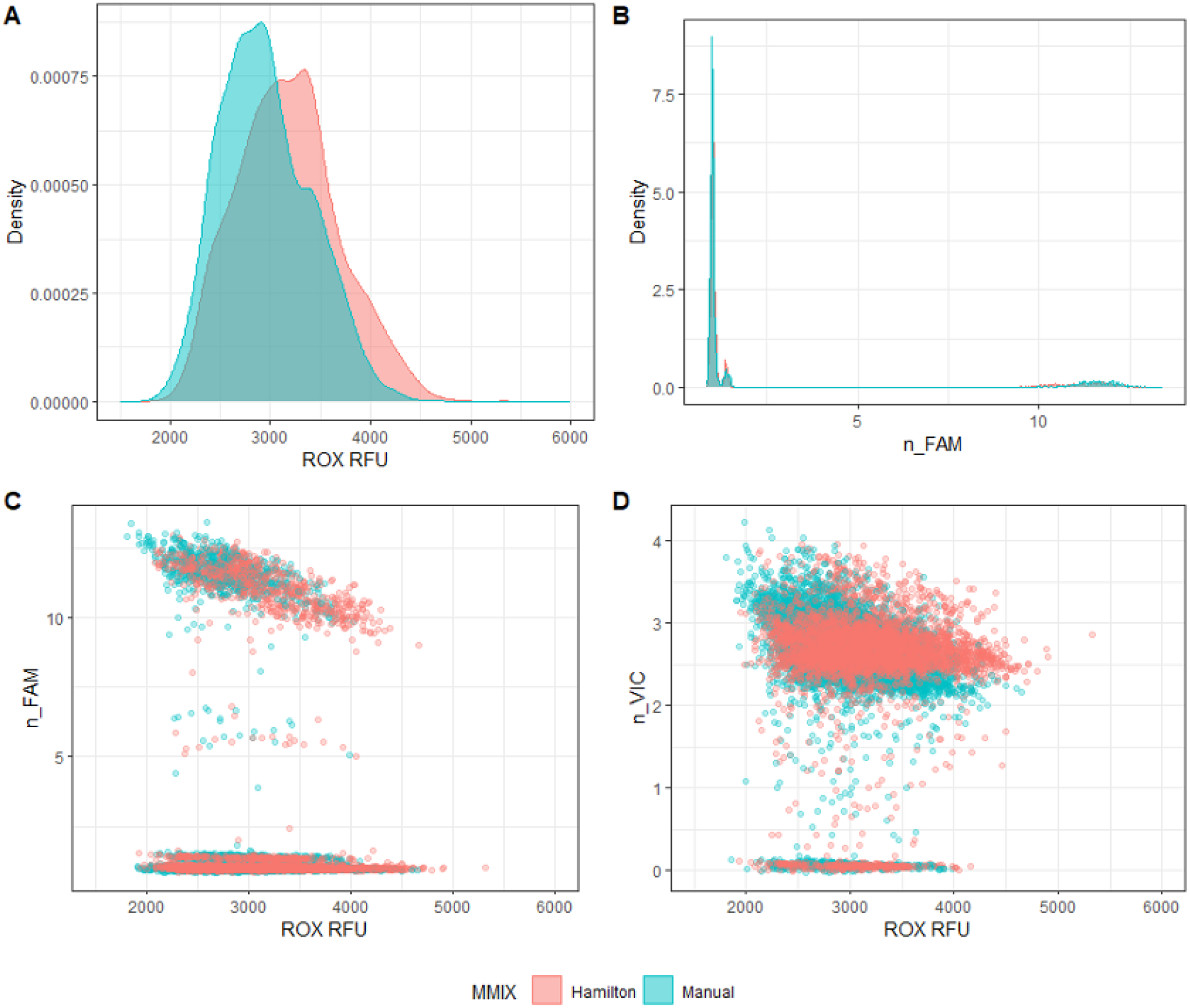
StarLet-Prepared MMIX compared with Manually-Prepared MMIX. End-point RFU data from PCR reactions using patient samples and control material used in routine clinical analysis. A) ROX RFU kernel distribution B) FAM RFU kernel distribution. C) Scatterplot n FAM vs ROX RFU. D) Scatterplot n VIC vs ROX RFU. Data are pooled from twelve 384-well arrays for both conditions.

**Table 10:** Cohen’s Kappa analysis of percent agreement.

| Class | % | Cohen's Kappa |
| --- | --- | --- |
| PPA | 98.71 | 0.95 |
| NPA | 98.52 |  |
| OPA | 98.55 |  |

**Table 11:**
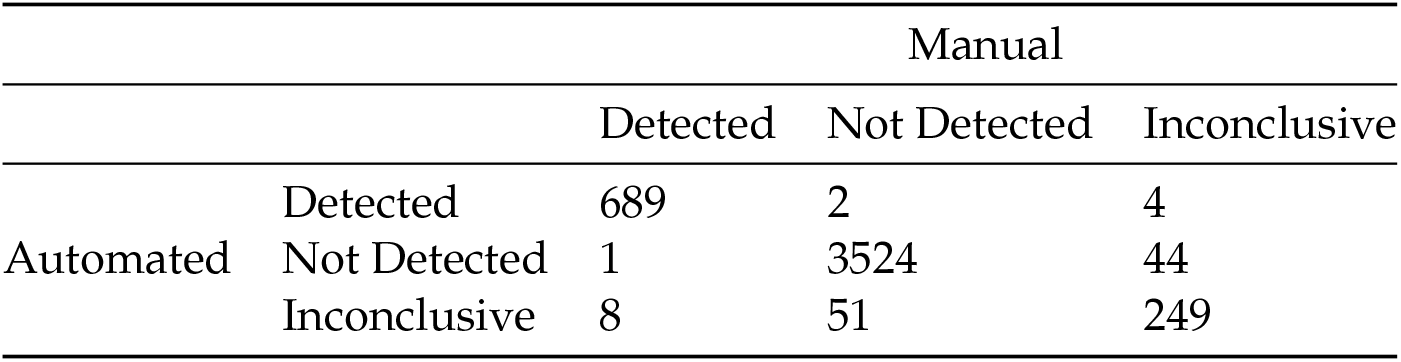
Confusion matrix of concordance results.

Together, the data collected during the validation demonstrate that the automated MMIX method gives results equivalent to trained and experienced operators. The method was therefore approved for routine operational use in the laboratory.

## 4 Discussion

In laboratories that provide routine molecular diagnostic services, technicians are often burdened with repetitive pipetting tasks, for instance transfer of sample from tube to microplate, or microplate re-formatting. Such tasks are prime candidates for automation, and barring unusual liquid classes, implementation tends to be straightforward. The preparation of MMIX, however, has been relatively neglected by process automators because MMIX reagents are expensive, reagents may require defrosting with a limited number of freeze-thaw cycles and a precise ratio of reagent volumes requires calculation ad hoc based on the amount of MMIX required. Taken together with the viscous or saponaceous nature of some MMIX reagents, the automation of MMIX preparation can present acute challenges to automation developers.

Our work on the development and validation of methods to automate MMIX preparation at a high-throughput molecular diagnostic facility resulted in significant savings to the laboratory through dead volume minimisation, avoidance of common errors, and the reduction of operator hands-on time. Due to equipment availability we took a two-pronged approach to development and validation of automation platforms.

We began development of an automated process using the SPT LabTech Dragonfly Discovery platform. The Dragonfly employs positive displacement syringes and with only minimal effort in optimisation of dispense and aspiration speed, we could achieve accurate dispense performance. However, with the Dragonfly we encountered a shift in the fluorophore signal distribution, specifically a marked reduction in ROX RFU. We hypothesised that this was because the carboxy-X-rhodamine fluorophore precipitated out of solution, aggregated, and adsorbed to the surface of the reservoirs. In support of this hypothesis, introduction of Tween-20 surfactant ameliorated ROX RFU to expected levels, perhaps through helping keep the carboxy-X-rhodamine fluorophore in solution. In addition, the Dragonfly did not have a built in sample or reagent tracking database, nor liquid level detection, hence a regulated operational environment would also require engineering or process controls to minimise the risk of operator error. To explore a process control we developed a spectroscopic quality control procedure using a BMG LabTech PheraSTAR platereader. We demonstrated that a spectroscopic method could determine if some common errors such as swapping the volumes of reagents had been made during MMIX preparation. On the other hand, the method was unable to detect small variations in Episcript volume. We note that it would be possible to implement dual spectroscopic and gravimetric quality control methods, which with development and honing of acceptable ranges could be used to detect abnormalities in MMIX preparation.

Key considerations in evaluating, developing and operationalising automated workflows are the operator experience of its use, compliance requirements and the need for process controls (Hawker, 2017). As the innate features of the Dragonfly platform did not include functions that could have implemented the required process controls without additional QC steps, we shifted development to the Hamilton StarLet platform, which could both provide liquid level detection, a pipette log audit trail, a customised user experience, and a tracking database.

We developed a fully bespoke method, using both custom and built in liquid classes, the logic of which was designed to maximise the opportunity for reagent recovery in the event of the method aborting. While all applications differ, and this exact method will not be directly applicable to other workflows, the underlying logic of it is re-usable. It is an example of how it is possible to minimise the risk of reagent wastage while providing consistent results. The liquid handling performance, after optimisation, gave distributions of fluorescent signals that very closely matched a skilled manual operator. We therefore proceeded to validate the automated method through a method transfer (Webster et al., 2005). The method passed acceptance criteria for the validation and was therefore introduced into routine use, where it was operated until the retirement of the SARS-CoV-2 screening service.

Further developments and improvements that could be explored include integration with Laboratory Information Management Systems (LIMS) and full process automation through collaborative robots. Indeed, laboratories of the future are trending towards greater levels of integration, automation (Abolhasani and Kumacheva, 2023) and even the implementation of artificial intelligence (Marescotti et al., 2022).

We hope that by making this work available, it provides an example to the laboratory automation community of development of liquid handling workflows for MMIX preparation, and the logic that can be used to minimise wastage. The automation of MMIX preparation is important to ensure consistency and quality in high throughput molecular diagnostic applications. Plans for scaling up operations in response to demand from a “Pathogen X” outbreak (Skyle, 2022) should factor in automation, and carefully consider the logic required to ensure demand and quality are both met.

## 5 Notes

The laboratory work was financially supported by the UK Department of Health and Social Care through the Test and Trace programme. The laboratory work was conducted at the Rosalind Franklin Laboratory (Royal Leamington Spa, UK). As all data were obtained from anonymised patient samples routinely collected as part of the Test and Trace programme, there were no prospective samples collected. The use of samples for PCR testing as described in this work is covered by the over-arching Privacy Notice issued by the UKHSA with the legal basis for processing such data being:

- GDPR Article 6(1)(e) – the processing is necessary for the performance of its official tasks carried out in the public interest in providing and managing a health service
- GDPR Article 9(2)(h) – the processing is necessary for the management of health/social care systems or services
- GDPR Article 9(2)(i) – the processing is necessary for reasons of public interest in the area of public health
- Data Protection Act 2018 – Schedule 1, Part 1, (2) (2) (f) – health or social care purposes

## 6 CRediT author statement

**G.F.** Conceptualisation, Methodology, Formal Analysis, Investigation, Writing - Original Draft; **G.H.** Conceptualisation, Methodology, Software; **A.S.** Methodology, Software, Validation; **S.L.** Conceptualisation, Methodology; **B.Y.** Validation, Investigation, Resources; **D.R.C** Validation, Investigation, Resources; **D.G.** Resources; **R.D.** Validation, Investigation; **P.E.R.-D.** Validation, Investigation, Project Administration, Supervision; **A.W.** Project Administration, Supervision; **A.D.B.** Project Administration, Supervision; **M.G.** Project Administration, Supervision; **B.P.** Project Administration; **S.G.** Project Administration; **D.P.F.** Data Curation, Writing - Original Draft, Project Administration, Supervision. All authors were responsible for review and editing of final manuscript.

## Data Availability

All data produced in the present study are available upon approval of a freedom of information request to the UK Health Security Agency (UKHSA).
Liquid Handler methods are freely available in a GitHub repository.

https://github.com/Donald-OMIX/Liquid_Handler_Methods

## 8. Acknowledgements

The authors thank the community that built up around the UK’s national COVID-19 testing program, which included both public and private sector organisations. We thank our friends and colleagues at LGC, Hamilton, SPT Labtech and BMG Labtech for their help and technical insights into the capabilities of their reagents and equipment. In particular we thank the Technology and Innovation Group at the Rosalind Franklin Laboratory whose support, leadership, and culture, created the right environment for work such as this to happen.

